# Factors affecting Cost-Related Medication Non-Adherence among US population with Cardiovascular Risk Factors

**DOI:** 10.1101/2024.03.11.24304136

**Authors:** Nikhila Gandrakota, Manju Ramakrishnan, Kavya Sudireddy, Megha K Shah

## Abstract

**Introduction:** In the US, cardiovascular diseases (CVD) are the leading cause of death and disability. Cost-related non-adherence (CRMN) can have serious consequences and worsen CVD outcomes. We examined the relationship between CVD risk factors and CRMN among US adults.

**Methods:** CDC’s 2019-2021 National Health Interview Survey (NHIS) data was used to examine CRMN among adults, categorized into three groups based on reported risk factors. We used chi-square tests, logistic regression to determine factors associated with CRMN.

**Results:** Among 49,464 participants, young, unmarried individuals, females, less educated, and participants from South had higher CRMN than older, married individuals, males, and those with higher education residing in the other regions. Current smokers and those with more CVD risk factors also had a higher CRMN than former smokers and never-smokers. Conversely, those aged 65 or older, with high-income, and excellent self-rated health had lower CRMN than younger participants, low-income families, and those with poor self-health. Public insurance and Medicaid participants had lower CRMN than uninsured (OR 0.13, 95% CI, 0.04-0.45, and OR 0.24, 95% CI, 0.15-0.36). Stratified regression analysis by disease status, i.e., diabetes, hypertension, and hyperlipidemia, revealed participants with high-income had lower odds of CRMN (OR 0.38, 95% CI 0.28-0.50; OR 0.39, 95% CI, 0.28-0.58; OR 0.37, 95% CI 0.27-0.51 respectively) than those with lower incomes.

**Conclusion:** Adults under 65 with more CVD risk factors are at higher risk of CRMN. Hence, robust prescription drug coverage and targeted interventions are necessary to lower CRMN in those with CVD risk factors.

## Introduction

Medication non-adherence tremendously impacts the healthcare system and costs the US healthcare system $500 billion per year.^1, 5, 6^. The costs incurred due to complications, hospitalizations, and further medical interventions resulting from the inability to take prescribed medication lead individuals into a medico-economic decline.^7, 8^ Several factors responsible include the high cost of medications, long-term treatment regimens, polypharmacy, insurance coverage, and lack of awareness.^2^ Chronic diseases like hypertension and diabetes often require the administration of complex multimodal medication regimens, and non-adherence can lead to disease exacerbation.^3, 4^ Lack of interventions to alleviate concerns about adherence to medications can lead to poor health outcomes.^9^

In the United States, non-adherence afflicts > 60% of patients diagnosed with cardiovascular disease (CVD), the leading cause of morbidity and mortality worldwide.^10, 11^ Additionally, the financial burden of CVD is projected to double and reach $1.1 trillion in 2035.^12^ At the forefront of this effect are risk factors of CVDs such as sociodemographic characteristics, tobacco smoking, physical inactivity, hypertension, diabetes, and dyslipidemia which if undermanaged, result in both the progression of the disease and excessive utilization of healthcare resources.^13^ Several other recent studies, demonstrated low adherence rates in 10% and 11.9% of subjects on anti-hypertensive and lipid-lowering agents respectively.^14^ In addition, Osborn et. al showed the significance of financial strain on non-adherence rates, highlighting the need to further evaluate cost-related medication non-adherence in the progression of CVDs.^15^ In our study, to better understand and identify the people at risk for cost related medication non-adherence (CRMN), we intend to explore the circumstances and characteristics of the population that can drive the same.

Through the analysis of a large nationally representative sample data of individuals, we describe sociodemographic, regional, and health-related factors influencing cost-related medication non-adherence among those with cardiovascular risk factors. The study aims to determine the underlying factors among adults with CVD risk factors to better inform what groups might be at risk for cost related nonadherence.

## Methods

We conducted a cross-sectional analysis of pooled data from the CDC’s National Health Interview Survey (NHIS) from years 2019-2021. The NHIS is an annual cross-sectional survey that collects information on the health and health-related behaviors of the U.S. population^16^ The National Center for Health Statistics (NCHS) oversees the NHIS data collection.

Adults aged 18 years and older who reported having at least one of three CVD risk factors: diabetes, hypertension, or hyperlipidemia, and who had received prescription medication in the past 12 months were included in the study. Participants were classified as having no, one, or more than one CVD risk factor based on their responses to whether they had ever been told they had diabetes, high blood pressure, or high cholesterol in the survey.

Cost-related Medication non-adherence (CRMN) was assessed from the questions which asked participants whether they have skipped medication doses to save money, taken less medicine to save money, or delayed filling a prescription to save money in the past 12 months. Participants were classified as having CRMN if they answered “yes” to any of these questions.

Sociodemographic characteristics of the respondents, including age, sex, education, marital status, income ratio to poverty, insurance status, obesity, smoking status, self-rated health status, and the number of adults and children in the household, were also assessed for the study. Chi-square tests were performed to assess the differences in cost-related non-adherence between the different categories.

Appropriate survey weights provided by the NHIS were applied to generate national estimates. Descriptive statistics were obtained using the percentage of cost-related non-adherence by different demographic and cardiovascular risk factor categories including age, sex, ethnicity, educational status, income ratio of poverty, and marital status. Chi-square tests were run to obtain p-values indicating the significance of the differences in cost-related non-adherence between the different risk factor categories. Logistic regression was performed to examine the association between CVD risk factors and cost-related medication non-adherence, adjusting for sociodemographic variables. Further, multivariate logistic regression analyses were also conducted by disease condition.

All statistical analyses were performed using SAS software, version 9.4 (SAS Institute, North Carolina, US)—a p-value less than 0.05% was considered significant.

## Results

### Demographics (Table 1)

In our study among a total of 49,464 participants (N weighted= 143,685,241) in the sample, we examined the proportion of cost-related medication non-adherence among various demographic groups based on the CVD risk factor status which is shown in Table 1. We found that 25.37% had no CVD risk factors, 41.93% had one risk factor and 32.79% had more than one risk factor.

**Table 1:**
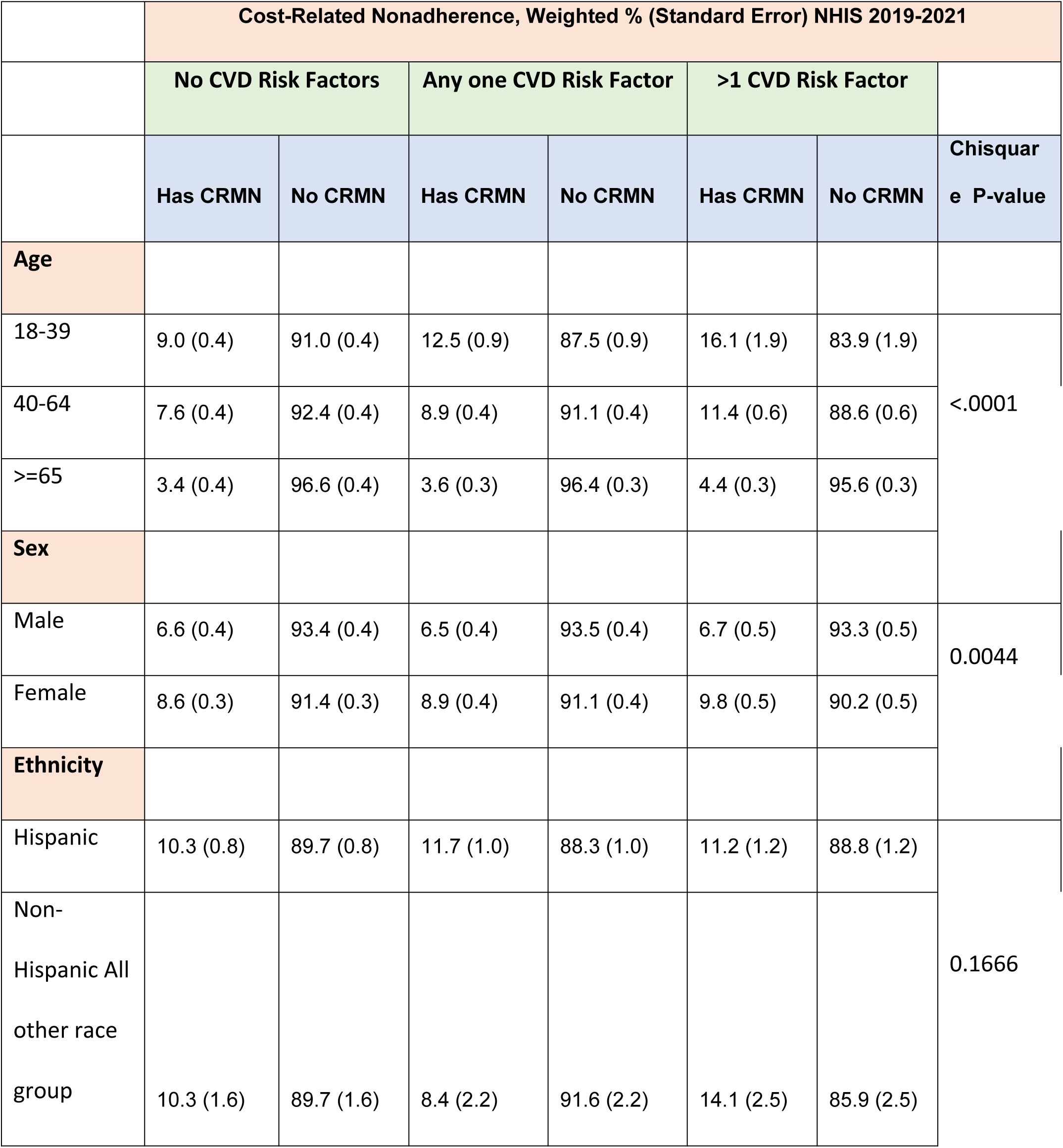
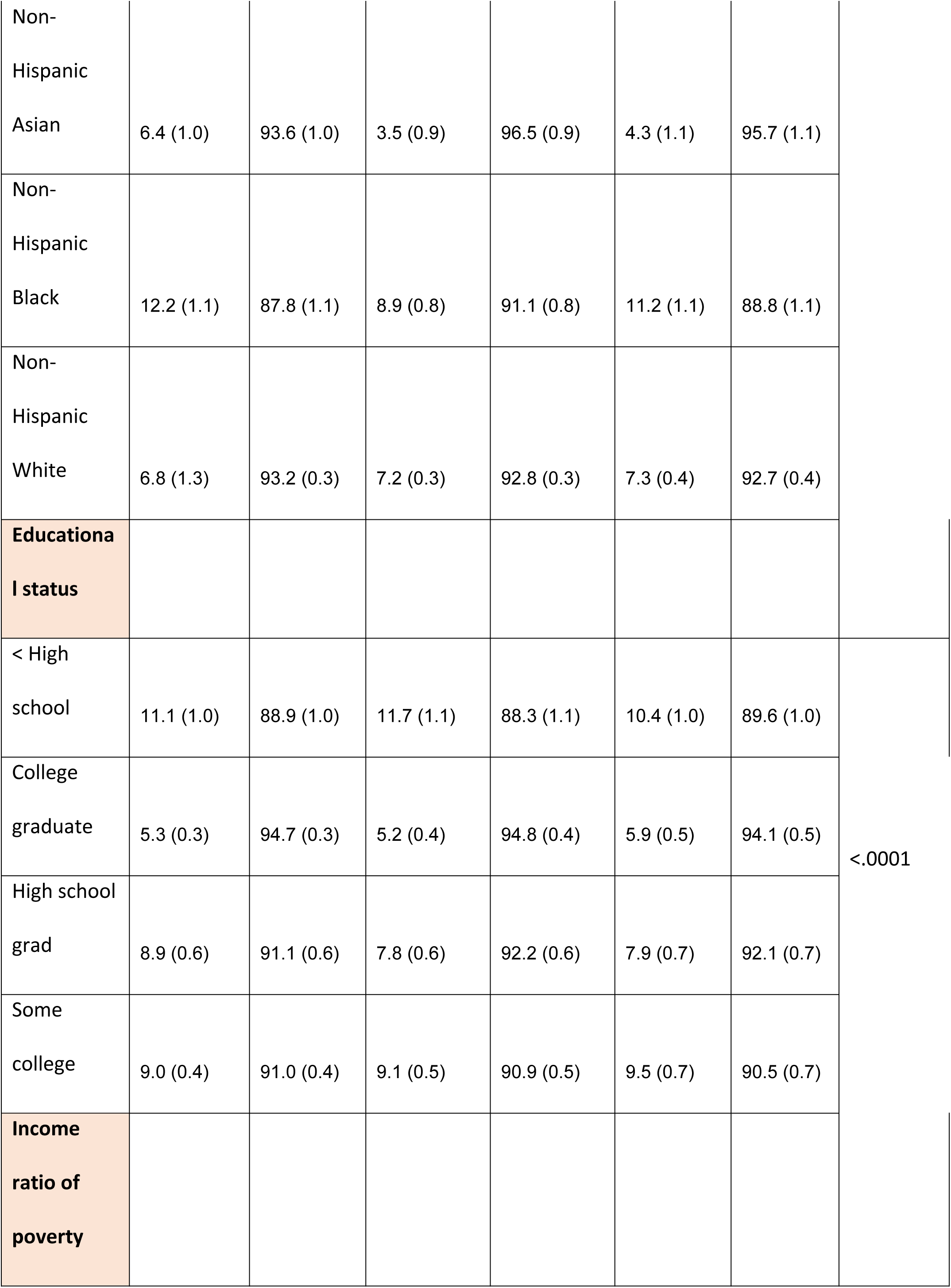
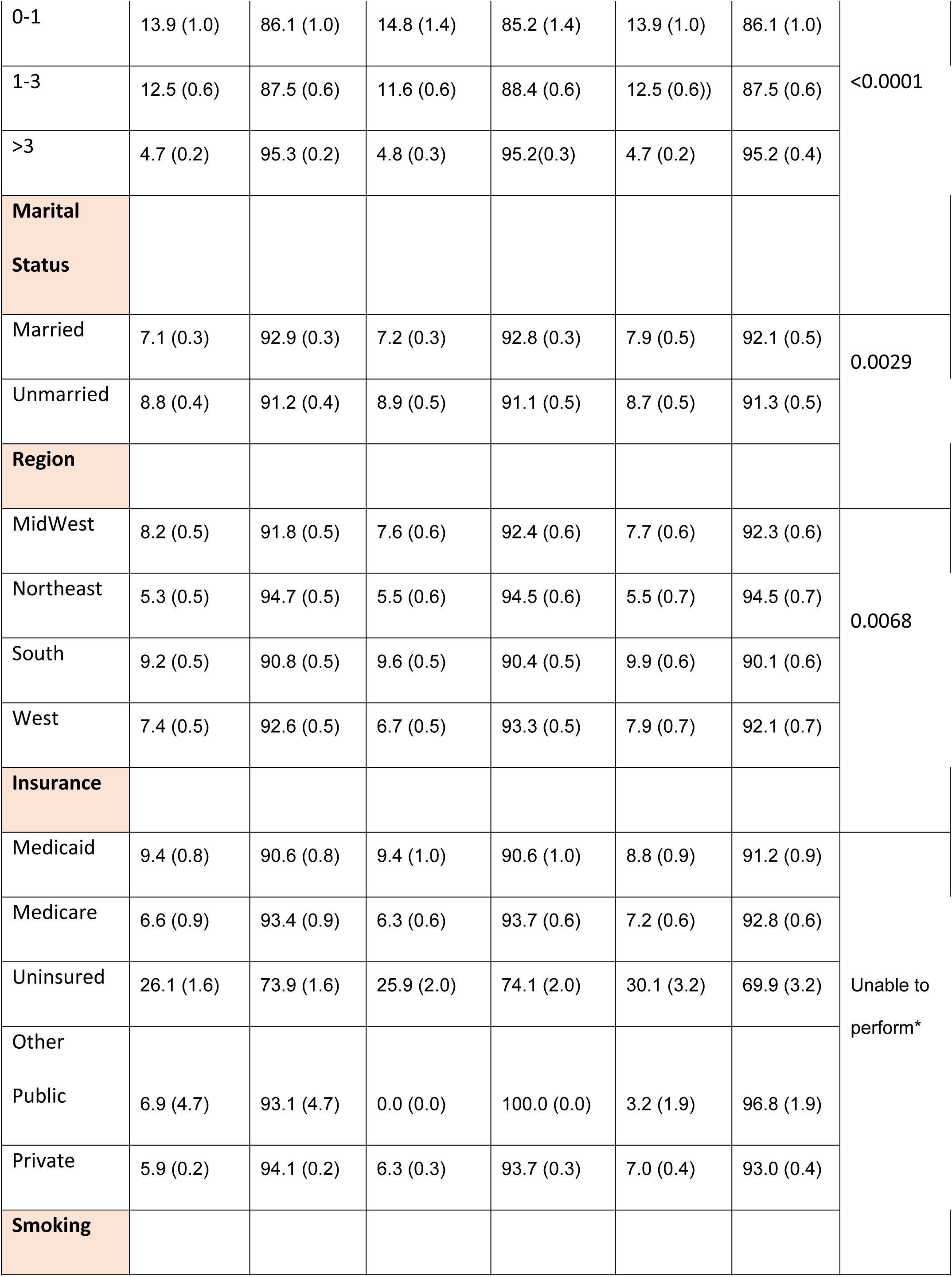
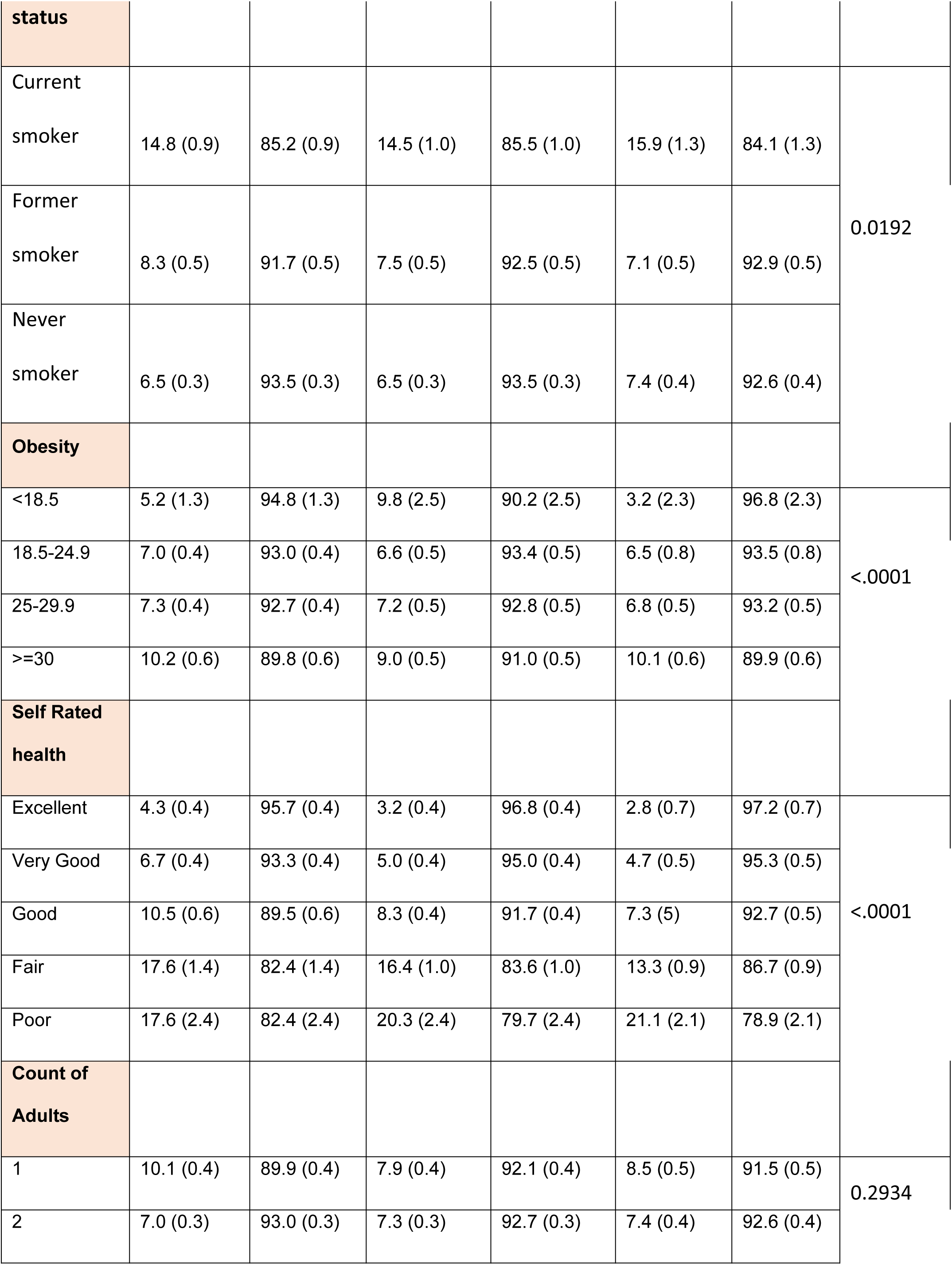
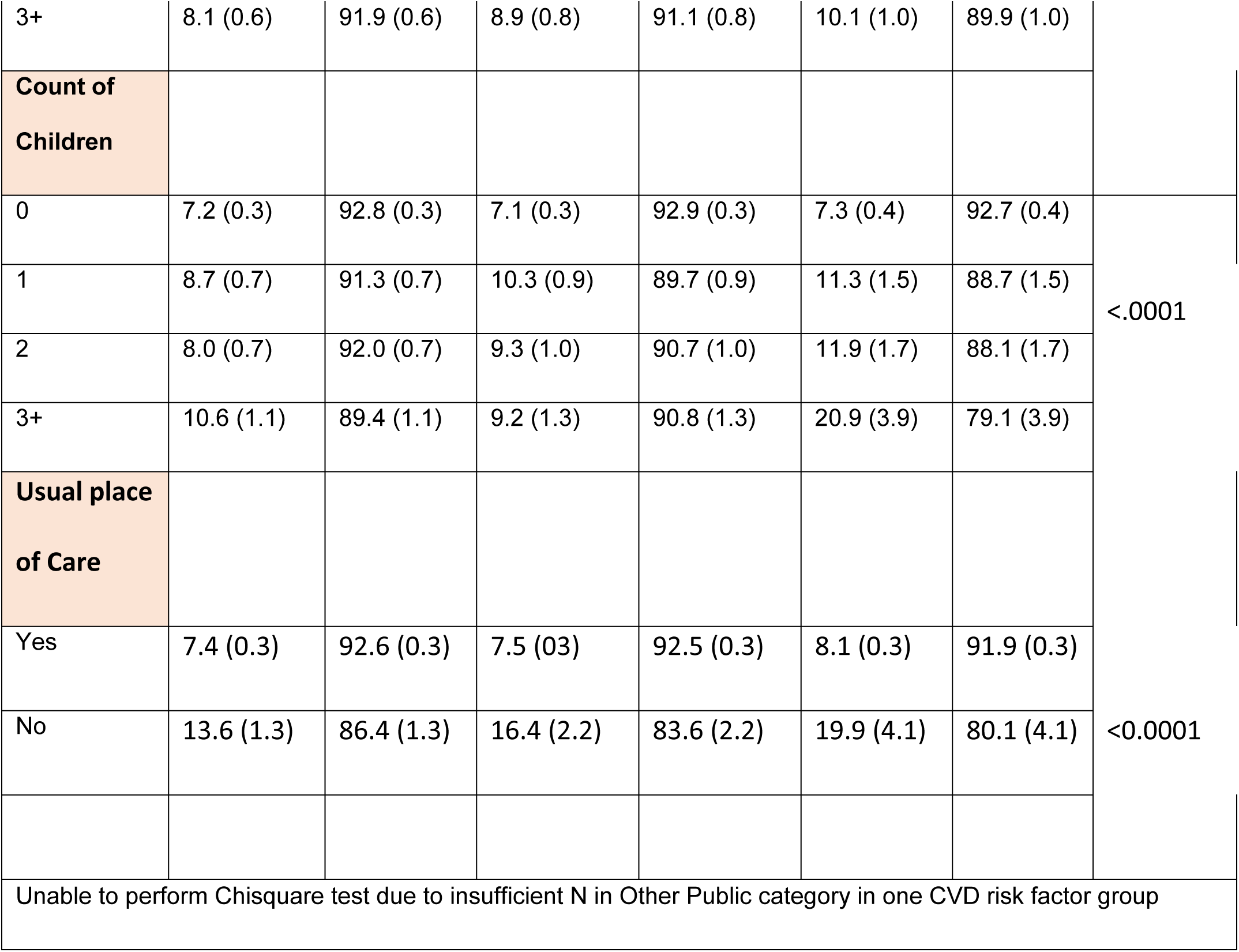
Characteristics of the study population with and without CRMN by risk factor status, (N=143685241) (n=49,464)

In participants with a history of more than one CVD risk factor, 16.1% among those who were aged between 18-39 years demonstrated CRMN while only 4.4% of the elderly population with similar CVD risk factor profile aged 65 years and above were non-adherent to medications. Patients with no CVD risk factors demonstrated a lower proportion of individuals exhibiting CRMN with 9% who fall within the age group of 18-39 years while 7.6% and 3.4% are between 40-65 years of age and over 65 years respectively. The differences in proportions were statistically significant (p<0.0001).

Females had a notably higher proportion of non-adherence irrespective of their risk factor status. Specifically, 8.6% of females with no CVD risk factors and 8.9%, and 9.8% with one and more than one CVD risk factor respectively compared to 6.6% of males with no CVD risk factors, 6.5% with one risk factor, and 6.7% with more than one CVD risk factor who showed non-adherence.

The level of education had a significant impact on the prevalence of CRMN among participants. Individuals with less than a high school level of education had a higher proportion of non-adherence than college graduates, with 11.1%, 11.7%, and 10.4% among those with no risk factors, one risk factor, and more than one risk factor, respectively (p=0.0001). Married respondents had lower non-adherence rates than unmarried respondents, with rates of 7.1%, 7.2%, and 7.9% for married patients with no risk factors, one risk factor, and more than one risk factor, respectively. Higher proportions of unmarried individuals were prone to be non-adherent. 8.8% of the unmarried respondents with no CVD risk factors displayed CRMN, with similar percentages of 8.9%, and 8.7% in individuals with one and more than one CVD risk factor correspondingly (p=0.0029).

The South exhibited highest non-adherence in patients with no risk factor, one, and multiple CVD risk factors, at 9.2%, 9.6%, and 9.9%, respectively compared to all other regions across the country. These regional differences across the country were statistically significant (p=0.0068). Furthermore, the results also showed that a higher prevalence of CRMN was observed in uninsured patients across all risk factor groups with 26.1%, 25.9%, 30.1% among those with no risk factors, one risk factor and more than one risk factors respectively.

Among respondents with more than one CVD risk factor and BMI greater than 30 there was a significant proportion (p< 0.0001) who were not adherent to medications (10.1%) Respondents who rated their health as fair or poor had higher non-adherence (p<0.0001) across all risk factor groups. Respondents with three or more children were significantly more likely to be non-adherent than those with no children (p<0.0001) across all risk factor groups. In addition, there was a higher prevalence of non-adherence in individuals who did not have an established place of care across all three risk factor groups (13.6% with no risk, 16.4% with one risk factor and 19.9% with more than one risk factor)

### Adjusted odds ratio of cost-related medication non-adherence (Table 2)

In adjusted models, participants who were >65 years of age had lower odds of cost-related medication nonadherence compared to 18-39 years across all three risk factor groups (Odds ratio of 0.31, 0.24 and 0.22 in no CVD risk factor, any one and more than one CVD risk factor groups respectively). Females with more than one CVD risk factor were 1.59 times more likely non-adherent than males (95% CI, 1.2-1.95). This was similar to the odds in the other two risk factor groups with 1.48 times in female participants with any one CVD risk factor (95% CI, 1.25-1.75) and 1.48 times in those without CVD risk factors (95% CI 1.24, 1.76). There were no statistically significant associations between medication adherence and the various race/ethnicity among all three of CRMN groups.

**Table 2:** Factors affecting CRMN by CVD Risk factor status: Results from multivariate logistic regression model.

|  | Cost-Related Nonadherence, OR (95% CI) |  |  |
| --- | --- | --- | --- |
|  | No CVD Risk Factors | Any 1 CVD Risk Factor | >1 CVD Risk Factor |
| **Age** |  |  |  |
| 18-39 | Reference | Reference | Reference |
| 40-64 | **0.77 (0.65, 0.91)** | **0.70 (0.56, 0.87)** | **0.63 (0.44, 0.90)** |
| >=65 | **0.31 (0.22, 0.43)** | **0.24 (0.17, 0.33)** | **0.22 (0.14, 0.34)** |
| **Sex** |  |  |  |
| Male | Reference | Reference | Reference |
| Female | **1.48 (1.24, 1.76)** | **1.48 (1.25, 1.75)** | **1.59 (1.28, 1.98)** |
| **Ethnicity** |  |  |  |
| Non-Hispanic White | Reference | Reference | Reference |
| Non-Hispanic Black | **1.47 (1.14, 1.90)** | 0.84 (0.65, 1.10) | 1.17 (0.87, 1.56) |
| Non-Hispanic Asian | 1.20 (0.84, 1.70) | 0.68 (0.39, 1.18) | 0.60 (0.33, 1.09) |
| Non-Hispanic All other race group | 1.09 (0.76, 1.58) | 0.56 (0.30, 1.04) | 1.28 (0.79, 2.08) |
| Hispanic | 1.08 (0.85, 1.36) | 0.99 (0.76, 1.28) | 0.91 (0.65, 1.27) |
| **Educational status** |  |  |  |
| < High school | Reference | Reference | Reference |
| High school graduate | 1.12 (0.86, 1.47) | 0.92 (0.68, 1.24) | 1.09 (0.80, 1.49) |
| Some college | **1.43 (1.11, 1.83)** | 1.27 (0.94, 1.71) | **1.51 (1.11, 2.06)** |
| College graduate | 1.29 (0.98, 1.69) | 1.16 (0.84, 1.59) | **1.44 (1.01, 2.05)** |
| **Income ratio to poverty** |  |  |  |
| <1 | Reference | Reference | Reference |
| 1-1.99 | 0.98 (0.79, 1.23) | 0.96 (0.70, 1.30) | 1.13 (0.84, 1.53) |
| >2 | **0.49 (0.37, 0.64)** | **0.46 (0.33, 0.66)** | **0.52 (0.36, 0.76)** |
| **Marital Status** |  |  |  |
| Unmarried | **Reference** | **Reference** | **Reference** |
| Married | 1.11 (0.90, 1.38) | 1.04 (0.81, 1.34) | 1.34 (0.99, 1.80) |
| **Region** |  |  |  |
| Northeast | **Reference** | **Reference** | **Reference** |
| MidWest | **1.47 (1.18, 1.84)** | 1.09 (0.82, 1.46) | 1.35 (0.95, 1.92) |
| South | **1.41 (1.12, 1.77)** | **1.32 (1.01, 1.72)** | **1.53 (1.09, 2.15)** |
| West | **1.39 (1.10, 1.77)** | 1.03 (0.76, 1.39) | 1.35 (0.93, 1.96) |
| **Insurance** |  |  |  |
| Uninsured | **Reference** | **Reference** | **Reference** |
| Private | **0.31 (0.25, 0.39)** | **0.38 (0.28, 0.50)** | **0.39 (0.26, 0.59)** |
| Medicaid | **0.24 (0.18, 0.32)** | **0.24 (0.17, 0.35)** | **0.24 (0.15, 0.36)** |
| Medicare | **0.44 (0.29, 0.67)** | **0.48 (0.32, 0.71)** | **0.59 (0.38, 0.91)** |
| Other Public* | 0.41 (0.10, 1.62) | **<0.001 (<0.001, <0.001)\*** | **0.13 (0.04, 0.45)** |
| **Smoking status** |  |  |  |
| Never smoker | **Reference** | **Reference** | **Reference** |
| Former smoker | **1.48 (1.23, 1.77)** | **1.38 (1.12, 1.69)** | 1.23 (0.99, 1.51) |
| Current smoker | **1.73 (1.42, 2.10)** | **1.78 (1.40, 2.26)** | **1.82 (1.40, 2.37)** |
| **Obesity** |  |  |  |
| <18.5 | **Reference** | **Reference** | **Reference** |
| 18.5-24.9 | 1.66 (0.93, 2.95) | 1.39 (0.70, 2.76) | 2.79 (0.53, 14.58) |
| 25-29.9 | 1.58 (0.89, 2.81) | 1.57 (0.78, 3.16) | 2.57 (0.49, 13.53) |
| >=30 | 1.72 (0.96, 3.07) | 1.47 (0.73, 2.95) | 2.76 (0.52, 14.58) |
| **Self Rated Health** |  |  |  |
| Good | **Reference** | **Reference** | **Reference** |
| Excellent | **0.46 (0.36, 0.59)** | **0.45 (0.32, 0.63)** | **0.47 (0.27, 0.82)** |
| Very good | **0.70 (0.58, 0.84)** | **0.67 (0.54, 0.84)** | **0.70 (0.51, 0.95)** |
| Fair | **1.90 (1.48, 2.44)** | **1.96 (1.60, 2.41)** | **1.52 (1.20, 1.92)** |
| Poor | **1.88 (1.26, 2.79)** | **2.61 (1.85, 3.68)** | **2.94 (2.19, 3.96)** |
| **Count of Adults in the Household** |  |  |  |
| 1 | **Reference** | **Reference** | **Reference** |
| 2 | **0.76 (0.62, 0.94)** | 1.06 (0.83, 1.36) | 0.80 (0.59, 1.08) |
| 3+ | 0.84 (0.67, 1.04) | 1.07 (0.79, 1.44) | 0.95 (0.69, 1.31) |
| **Count of Children in the Household** |  |  |  |
| 0 | **Reference** | **Reference** | **Reference** |
| 1 | 1.12 (0.89, 1.42) | 1.03 (0.81, 1.31) | 0.98 (0.69, 1.39) |
| 2 | 0.90 (0.73, 1.12) | 0.88 (0.64, 1.20) | 0.89 (0.60, 1.34) |
| 3+ | 1.02 (0.79, 1.32) | **0.63 (0.42, 0.94)** | 1.44 (0.89, 2.34) |
| **Usual Place** |  |  |  |
| Yes | **Reference** | **Reference** | **Reference** |
| No | 1.32 (1.02, 1.71) | 1.43 (0.99, 2.08) | 1.38 (0.83, 2.29) |
| *Unable to determine an accurate estimate as the analysis produced a considerably lower odds ratio and corresponding confidence interval |  |  |  |

Respondents who attended some college and had more than one CVD risk factor were 1.51 times more likely to be nonadherent (95% CI, 1.11-2.06) while those who had no risk factors were 1.43 times more likely to be nonadherent compared to people without high school education (95% CI, 1.11-1.83).

High-income families (ratio more than or equal to 2) has a lower odds of CRMN compared to low-income families (ratio less than 1) with an odds of 0.49 (95% CI, 0.37-0.64), 0.46 (95% CI, 0.33-0.66), and 0.52 (95% CI, 0.36-0.76) in no CVD risk factor, any one CVD risk factor and more than one CVD risk factor groups respectively. The odds of CRMN in people with any insurance were significantly less than the uninsured, with those on Medicaid who had the lowest odds of 0.24 in respondents across all three groups [no risk factors (95% CI, 0.18-0.32), any one (95% CI, 0.17-0.35), and more than one CVD risk factor (95% CI, 0.15-0.36) groups]. Among patients who had more than one CVD risk factor, participants from the South had the highest odds and were 1.53 times more likely to be non-adherent compared to the Northeast (95% CI, 1.09-2.15).

Current smokers with multiple CVD risk factors were 1.82 times more likely to be non-adherent compared to those who never smoked (95% CI, 1.40-2.37) with similar odds in the no CVD risk factor and respondents with any one CVD risk factor.. In the same group, patients who had rated their health as poor were 2.94 times more likely to demonstrate higher CRMN than those who reported their health to be good (95% CI, 2.19-3.96) while those who rated their health as very good and excellent reported less CRMN (corresponding OR : 0.70, 95% CI, 0.51-0.95 and 0.47, 95% CI, 0.27-0.82).

### Association based on disease status (Table 3)

After controlling for age, sex, ethnicity, educational status, income ratio to poverty, marital status, region, insurance, participants were stratified based on the disease status, which included hypertension, diabetes and dyslipidemia. Patients who were more than 65 years of age with other public insurance or Medicaid had lower odds of non-adherence irrespective of whether they had hypertension, diabetes, hyperlipidemia. Similarly, respondents from high income households also demonstrated lower odds of non-adherence across all the disease groups with odd ratio of 0.38 (95% CI, 0.28-0.50) for hypertension, 0.39(95% CI 0.28-0.58) for diabetes, and 0.37 (95% CI, 0.27-0.51) for hyperlipidemia. CRMN, when compared to non-Hispanic white patients, was significantly less likely in non-Hispanic Asians who had hypertension (OR 0.55, 95% CI, 0.34-0.89) and diabetes (OR 0.35, 95% CI, 0.17-0.69). Further, females and those from the South had significantly higher odds of CRMN regardless of the disease category.

**Table 3:**
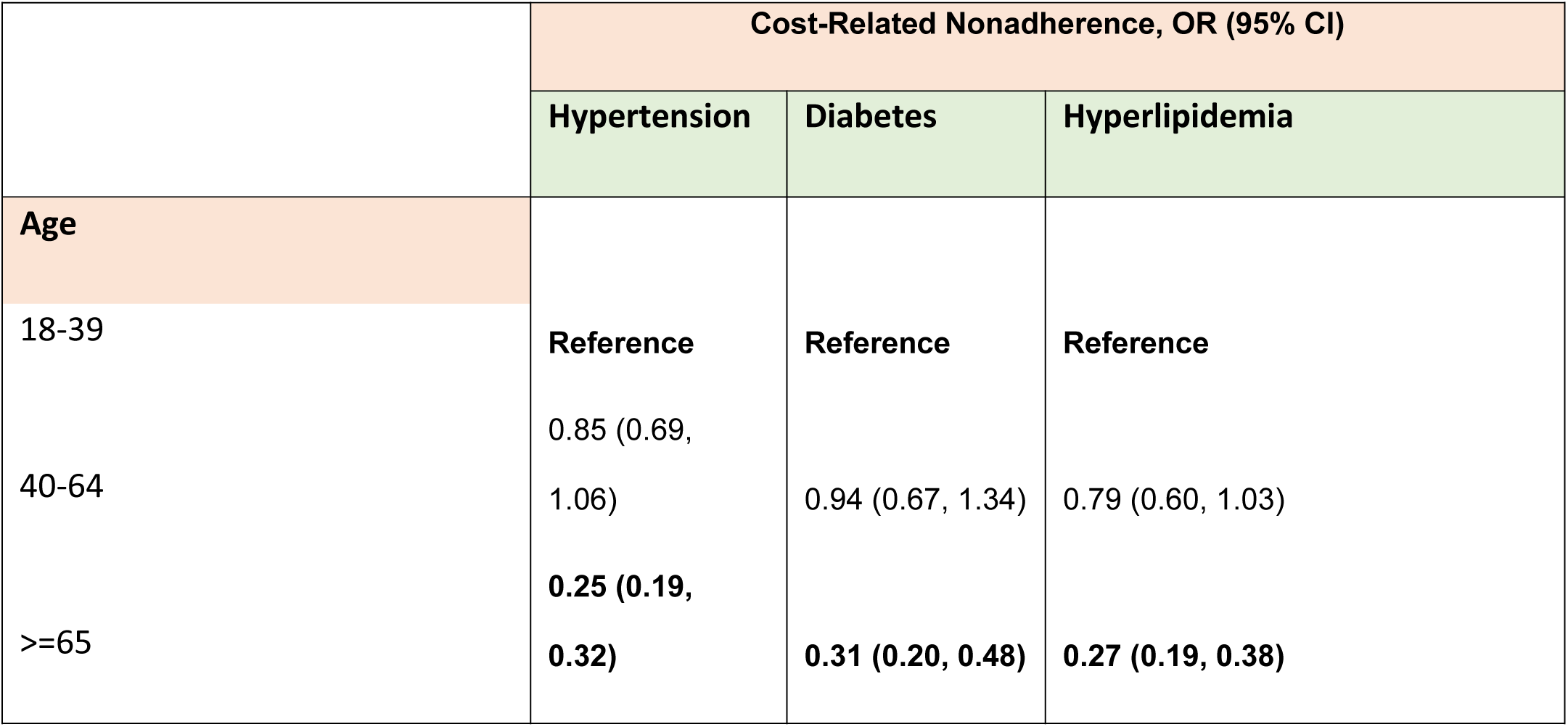
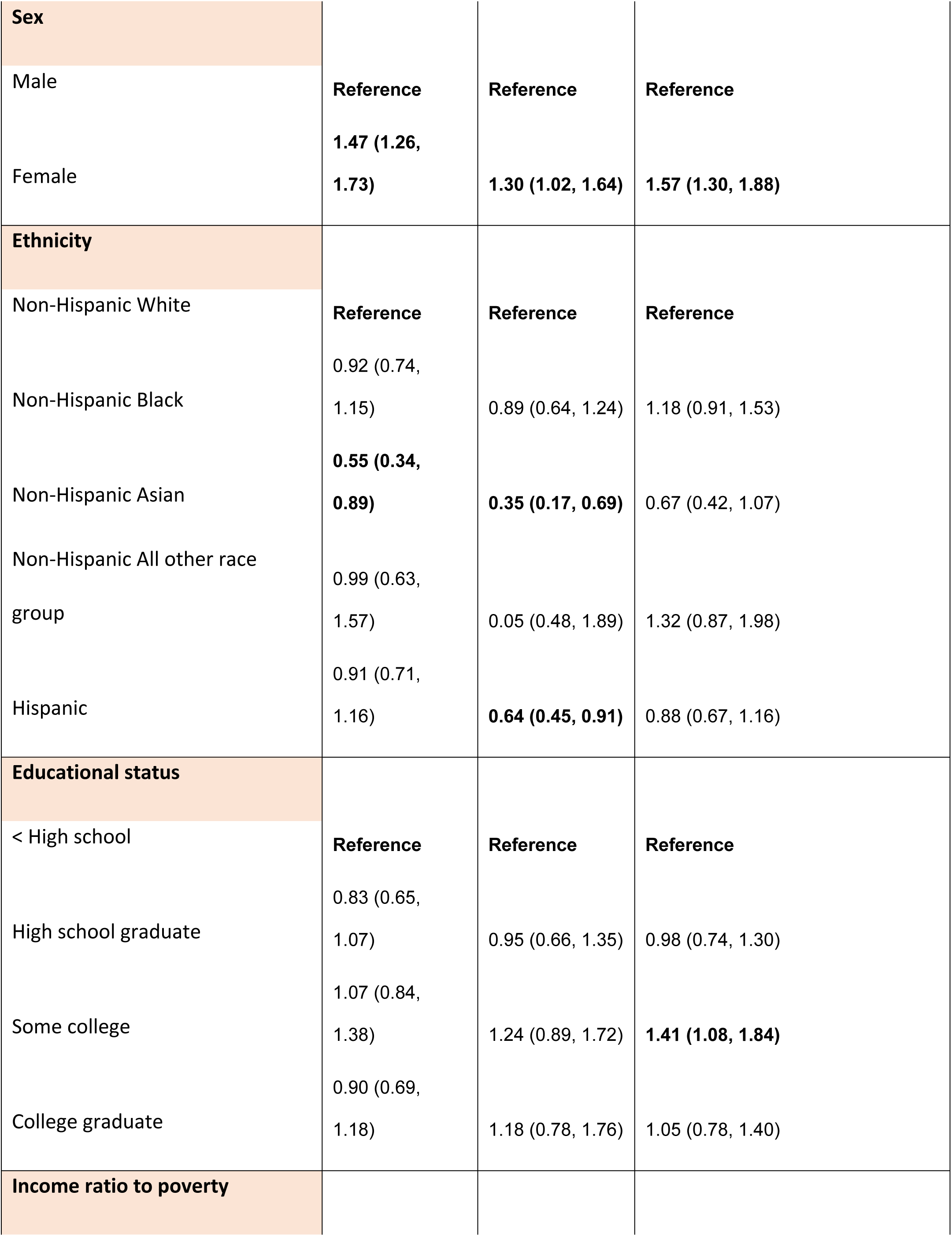
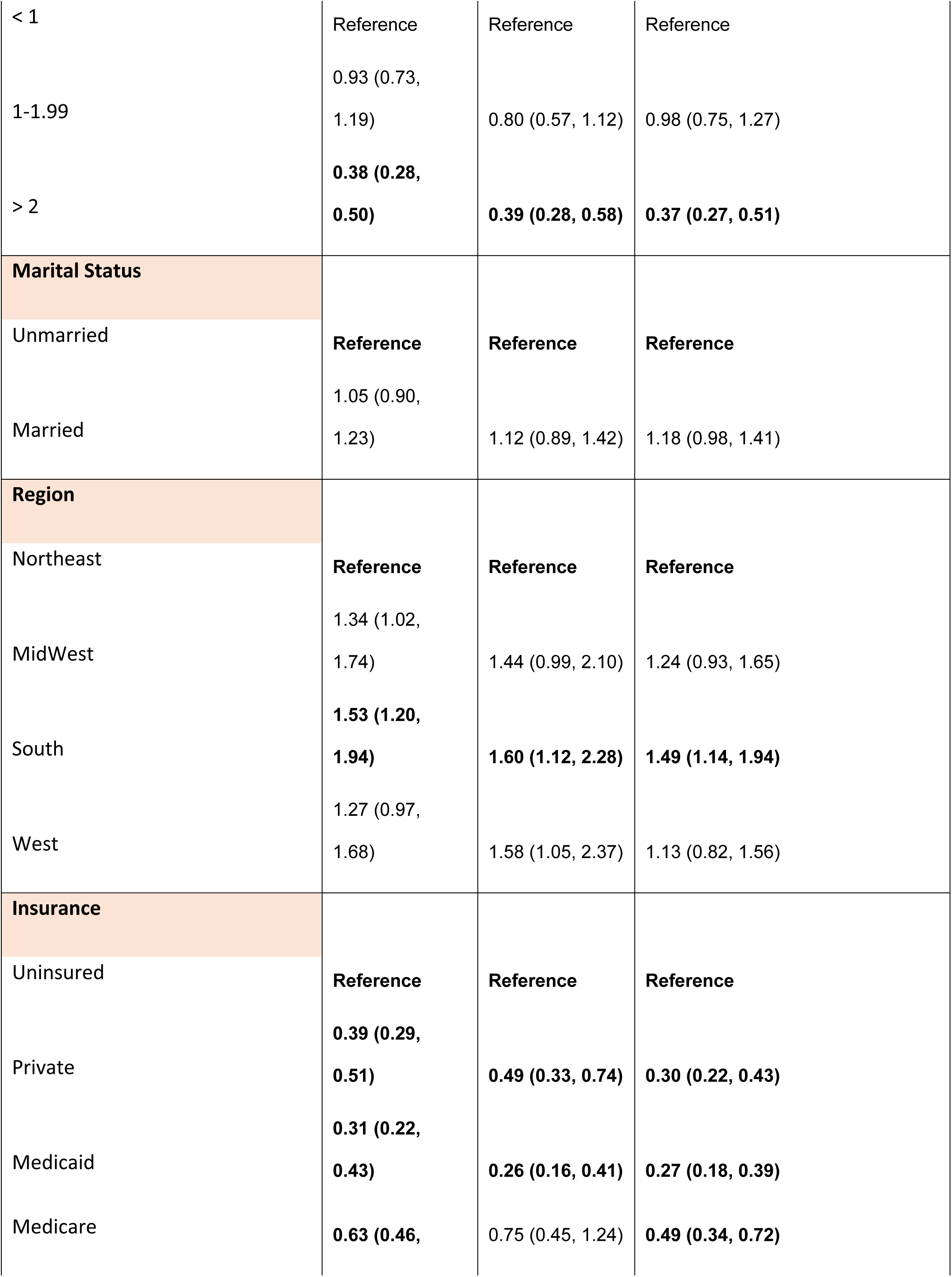
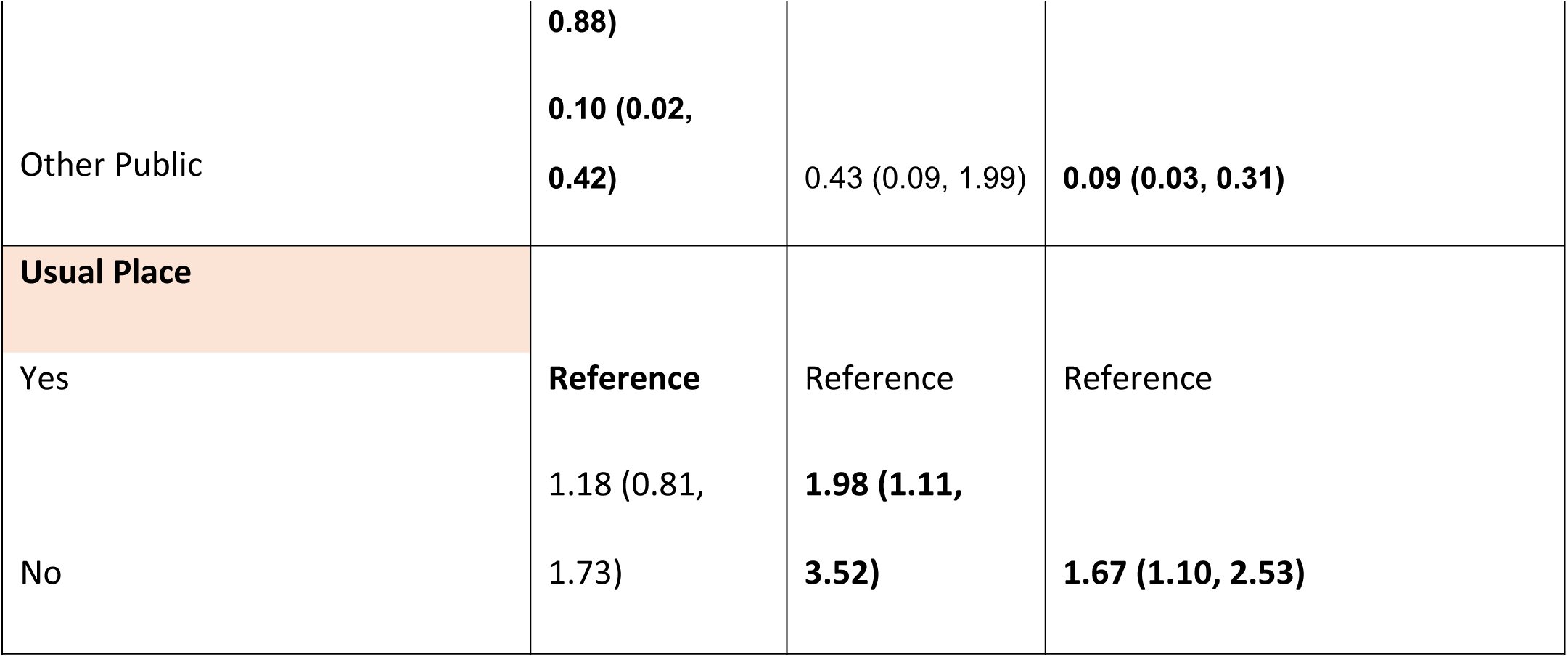
Factors affecting CRMN by Disease status: Results from multivariate logistic regression model.

## Discussion

Overall, CRMN varies by sociodemographic and disease status among adults living in the US. Young respondents, unmarried individuals, females, those with less education, and participants in the South had a higher proportion of CRMN compared to older, married individuals, males, and those with higher education and residing in the other regions. Our results suggest that non-adherence was lowest in participants over 65 years of age among all three risk factor groups (3.4% in participants with no CVD risk factors, 3.6% in those with one CVD risk factor and 4.4% among individuals with more than one CVD risk factors). This could be explained by the rise in the number of comorbid diseases in older individuals, increasing health awareness with age, greater resource allocation towards medications, and better prescription drug benefits like the Medicare Part D program.^16, 17^

Our analysis also determined that females with more than one CVD risk factor had higher odds of being non-adherent compared to males (OR 1.59, 95% CI 1.28-1.98). Although previous evidence shows that women are more likely to seek care compared to men, some studies suggest that women as primary caregivers prioritize their family’s health over their own.^18–20^ Additionally, an increasing financial burden based on the number of dependent children could also significantly contribute towards lower medication adherence in females.^18, 21^ This highlights the importance of developing better healthcare policies for younger adults not covered by Medicaid and women to facilitate reducing the disparities that are associated with CRMN.^17^

Among all the risk factor groups, higher proportions of CRMN were found among respondents who received less than the high school level of education followed by those with some form of college education.^22^ The former may face significant strain with a higher chance of being unemployed or failing to maintain a steady income.^23^ Either situation can cause financial strain and affect their insurance status as adults under 65 are primarily dependent on their employer-based insurance thereby impacting their medication adherence. ^23–25^ In a cross-sectional analysis of the NHIS data between 2010-2015, Su, Chia-Ping et al. determined that people with non-standard jobs like free-lance work, temporary contract-based employment, and working in small establishments were more likely to be uninsured.^26^ Our findings suggest that a robust social safety net that includes drug benefits might help mitigate the risk of CRMN among low-income and lower education groups.

Studies have found that poor economic status and underlying disease were negatively associated with self-rated health. ^27, 28^ The inability to access health resources due to financial hardship aggravates psychological distress and causes deterioration of overall well-being, making it more challenging to afford medications. ^29^ This may explain our findings that respondents from high-income households were more likely to be adherent compared to lower-income families irrespective of their disease status or the risk factor groups. Our study also determined that, within all risk factor groups, participants with three or more children had significantly higher non-adherence rates than those with no children. Studies found that families with children and in particular, low-income families are unable to bear the burden of insurance and out-of-pocket healthcare costs, which prove detrimental to their health needs.^30, 31^

In our assessment, although ethnicity was not a notable contributing factor toward medication adherence, there were significant regional differences observed across the country. Individuals from the South with more than one CVD risk factor were 1.59 times more likely than those from the Northeast to experience CRMN. It is prudent to note that some of the states in the Southern region of the U.S. are among the few that have chosen not to expand their Medicaid. According to the Centers for Disease Control, the residents of the Medicaid non-expansion states were more likely to be uninsured as they are neither eligible for Medicaid nor for other subsidized insurance coverage plans if their income is in the low to moderate range which would potentially affect medication adherence among these populations. ^32, 33^ Besides insurance, the regional variation in adherence can also be affected by factors like polypharmacy and complex drug regimens involving taking multiple pills at different times of the day, poor access to pharmacies especially in rural areas, and higher out-of-pocket expenses in areas with predominantly low-income populations.^34^ These can be rectified by introducing better prescription practice guidelines involving generic fixed-dose drug combinations which will reduce the costs as well as the number of pills, increasing doorstep delivery of medications by switching to mail-order or online pharmacy practices.^35, 36^

Our results also revealed that CRMN was higher among current smokers compared to non-smokers. This may be directly linked to the greater expenditure of their annual income on tobacco in comparison to medication.^37^ Additionally, in those classified as obese with a BMI of greater than 30, CRMN was significantly higher than in those with lower BMIs. Previous studies propose that weight-based bias perpetuated by the community has possibly led to less desire and motivation to seek care by overweight or obese people. Similarly, physician perceptions that obese patients are less likely to be adherent can further reinforce the bias by influencing the management and treatment of these. Interestingly, an individual’s annual medical care costs increase significantly with an increase in the class of obesity, going from 68.4% in class 1 to 233.6% in class 3 which can be due to a greater number of comorbidities in obese patients. ^38^ With rising healthcare costs directly attributed to the struggle to afford medication, it is imperative to strategize and promote healthy habits. ^39^ Furthermore, Lee et. al demonstrated that lifestyle modifications including cessation of smoking and a weight-loss diet were statistically associated with medication adherence, indicating the importance of interventions such as motivational education programs. ^40^ These features are potential modifying factors that are attributable to CRMN.

## Strengths and Limitations

Our study is strengthened by the sample size. Further, NHIS data is nationally representative, and hence the estimates are generalizable to the US population. Our study is also the first to use nationally representative data to examine the prevalence and factors associated with CRMN among people with and without CVD risk factors. However there are a few limitations to the study. One of the reasons for lower response rates in 2020 and 2021 compared to 2019 were challenges during the COVID-19 pandemic, with restriction on face-to-face data collection and hesitancy of respondents to participate during the pandemic. Since the data is self-reported, it might have been subjected to recall bias, social desirability bias, and other measurement biases. Finally, the data is cross sectional, limiting its use for causal inference and to observe temporal relationships between various health factors.

## Conclusion

Our study revealed variation in CRMN among the US population with and without cardiovascular risk factors. Higher proportion of CRMN was observed in younger ages, females, those with less education, unmarried respondents, and participants in the South. Further younger participants and those with multiple CVD risk factors without Medicare and Medicaid had a higher proportion of non-adherence compared to the older population Our findings highlight the need for targeted interventions to address these underlying factors in order to reduce cost-related medication non-adherence. For example, like the clinical decision support system, further research can be performed to analyze the possibility of incorporating screening guidelines using electronic medical records systems to identify the risk of non-adherence based on factors like prescription refills in primary care practices. Additionally, there is a need to evaluate the efficiency of prior authorization checks which when excessive have contributed to delays in treatments as well as non-adherence due to the inconveniences people experience in obtaining the medications.^41, 42^ Another key point to examine is the capping of prices of other expensive medications as it was done in the case of insulin for Medicare Part D enrollees while also expanding this coverage to Medicaid enrollees. Furthermore, negotiating price controls with pharmaceutical companies, bulk purchasing, and expediting the introduction of generic medications could help mitigate CRMN.^43–45^ This could provide greater access and coverage of affordable prescription drugs. Our study demonstrates the need for such systemic and policy changes to eliminate cost barriers to medication among all age groups.

## Data Availability

Data is openly available through CDC

## Abbreviations

NHIS: National Health Interview Survey
CDC: Centers for Disease Control and Prevention
NCHS: National Center for Health Statistics
CVD: Cardiovascular Disease

## Acknowledgments

None

## Author Contributions

NG had full access to all of the data in the study and is responsible for the integrity of the data and the accuracy of the data analysis. NG and MKS contributed to study design and conception, data interpretation and analysis, drafting of manuscript and final approval. MR and KS contributed to data interpretation, drafting of manuscript, critical review of manuscript and final approval.

## Notes

### Competing Interest Statement

The authors have declared no competing interest.

### Clinical Trial

NA

### Funding Statement

MKS is supported in part by NIMHD (K23 MD015088-04) and Emory School of Medicine Doris Duke Charitable Foundation COVID-19 Fund to Retain Clinical Scientists and the Georgia CTSA (UL1-TR002378)

### Author Declarations

Publicly available dataset.

